# Point-of-Care Biomarker Assay for Rapid Multiplexed Detection of CRP and IP-10

**DOI:** 10.1101/2023.05.25.23290476

**Authors:** Claire S. Wilson, Bhavya Vashi, Pavol Genzor, Melissa K. Gregory, Jason Yau, Lauren Wolfe, Michael J. Lochhead, Chris Myatt, Kristen Pettrone, Paul W. Blair, Subramaniam Krishnan, Josh G. Chenoweth, Danielle V. Clark

## Abstract

Rapid and accurate measurements of immune protein markers are essential for diagnosis and treatment in all clinical settings. The recent pandemic has revealed a stark need for developing new tools and assays that could be rapidly used in diverse settings and provide useful information to clinicians. Here, we describe the development and test application of a novel one-step CRP/IP-10 duplex assay for the LightDeck platform capable of delivering reproducible and accurate measurements in under eight minutes. We used the optimized assay to measure CRP and IP-10 levels in human blood and serum samples from healthy, COVID-19-positive, and influenza-like illness (ILI) presenting patients. Our results agreed with previously published analyte levels and enabled us to make statistically significant comparisons relevant to multiple clinical parameters. Our duplex assay is a simple and powerful tool for aiding diagnostic decisions in diverse settings.

## Introduction

SARS-CoV-2 infection is characterized by rapid viral replication and relatively mild symptoms (1-5 days) followed by a dysregulated immune response that can rapidly progress to severe symptoms and critical complications.^1^ Clinical parameters, including patient demographics, vital signs, comorbidities, radiographic images, and the host immune biomarker profile, are critical in understanding the pathophysiology and disease trajectory of COVID-19. The COVID-19 pandemic has had an unprecedented burden on hospital resources, highlighting the need for rapid diagnostic testing to facilitate risk stratification and guide clinical management. Previous studies showed that host response biomarkers could be leveraged to assess the patient’s immune state and triage medical care.^2–5^ Therefore, we aimed to develop a duplex biomarker assay to enable rapid, on-site, quantitative detection of biomarkers in whole blood. In this study, we describe a biomarker assay utilizing C-reactive protein (CRP) and chemokine CXCL10 (IP-10), both of which have been linked to the length of hospital stay, ICU admission, mechanical ventilation, and mortality. ^5–8^ CRP is associated with inflammation in severe COVID-19 and has been used as a diagnostic marker to describe the severity and prognosis of COVID-19.^9^ Elevated levels of IP-10 have been linked to viral disease progression, severity, morbidity, and mortality in COVID-19.^5,10,11^ Combining these two proteins as diagnostic markers can offer improved accuracy in discriminating between different stages of disease progression in COVID-19.^12,13^ CRP is a commonly measured analyte during routine hospital blood studies; however, IP-10 information is usually not accessible to physicians. This is because IP-10 measurements require long (hours to days) and tedious laboratory processing including sample collection, processing, and analysis by ELISA or LUMINEX assays. These assays are costly and require specialized laboratory equipment and facilities, significantly limiting their use in most clinical settings. The LightDeck platform and our CRP/IP-10 duplex assay (Figure 1A, B) offer a cost-effective solution requiring minimal resources and results comparable to other state-of-the-art laboratory testing methods. The LightDeck technology is based on a disposable cartridge analyzed on a portable reader with a proven broad range of applications in diagnostics and detection. ^14–16^ The cartridge uses an excitation laser traveling through a plastic substrate generating an evanescent illumination field that is read by the camera (Figure 1C).^17^ Planar waveguide imaging technology has been demonstrated for use in HIV-1 diagnosis^18^ and detection of Kaposi’s Sarcoma-Associated Herpesvirus. ^14^ We believe this system will help fill the current need for a rapid, easy-to-use, field-portable system for real-time monitoring of blood biomarkers.^12^

**Figure 1.**
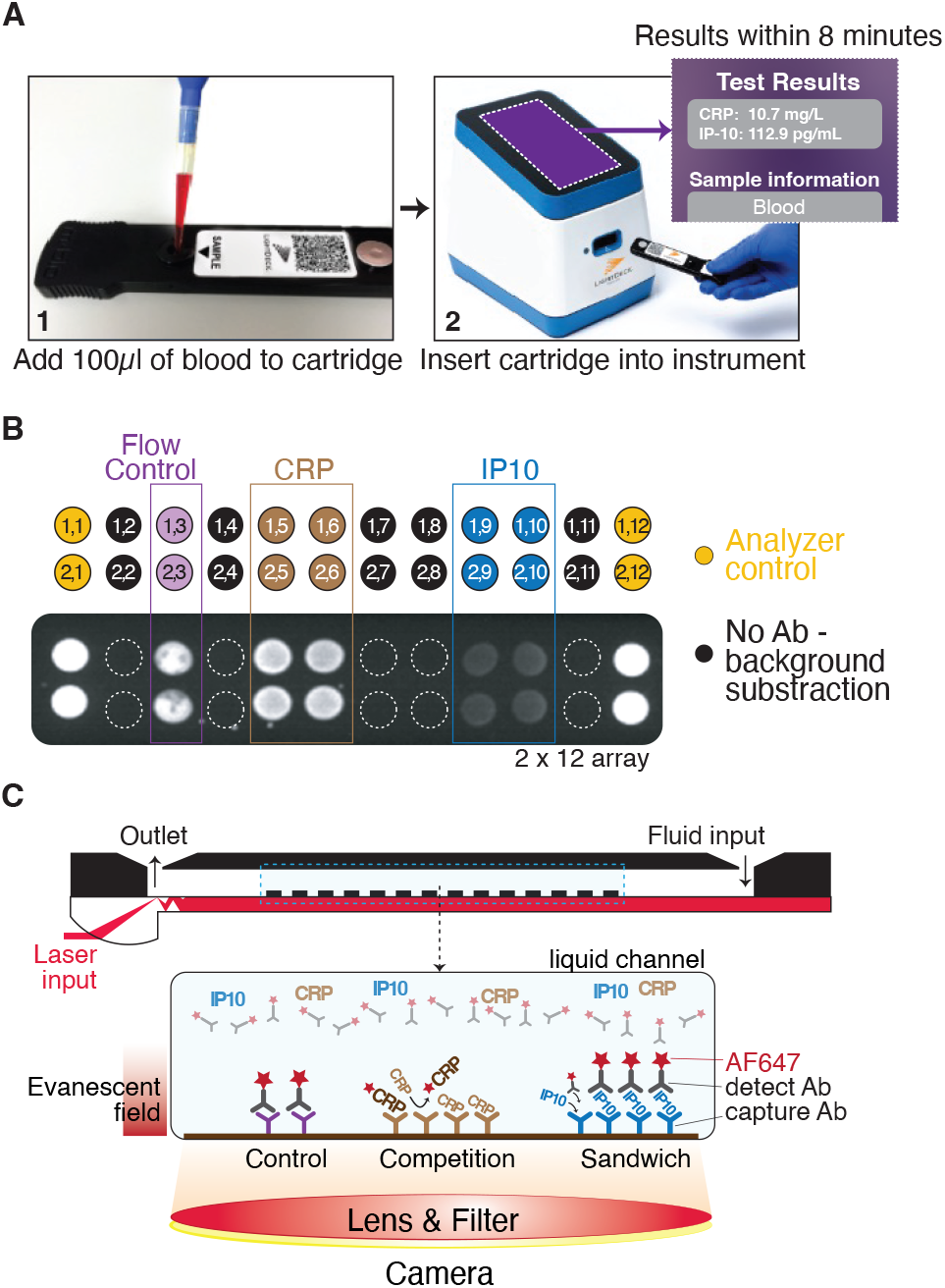
Lightdeck assay for rapid point-of-care quantification of CRP and IP-10. A) The LightDeck Platform test procedure is comprised of two simple steps and delivers results within 8 minutes. B) An array map and representative image of the CRP IP-10 Duplex test cartridge. C) A Cross-sectional view of the cartridge highligting the mechanistic aspects of the assay. CRP is measured in competitive assay with signal being inversly proportional to CRP concentration in the sample, while IP10 concentration is directly related to signal strength via antibody (Ab) sandwitch assay. Free flowing detect Ab does not generate any appreciable signal because the flourophore is ourside of the evanescent field.

## Materials and Methods

### Calibration and Limit of Detection (LoD)

Assay sensitivity was calibrated using pooled CRP-depleted human serum. The serum was spiked with recombinant proteins at seven different concentrations of each analyte, and three replicate measurements were carried out for each concentration. The Limit of Detection (LoD) was estimated using five serum samples containing low levels of CRP and IP-10, prepared at analyte concentrations surrounding the assumed LoD. The calculation of LoD is derived from the precision profile approach in the Clinical and Laboratory Standards Institute EP17-A2 Guideline.^19^

### Whole Blood Linearity

Assay linearity for CRP and IP-10 measurements was demonstrated using seven samples containing different levels of analytes. Samples were prepared by mixing a high-concentration K2EDTA whole blood sample with a low-concentration K2EDTA whole blood sample. In this case, the high sample was K2EDTA whole blood spiked with purified CRP at 150 mg/L and spiked with recombinant IP-10 at 2000 pg/mL, while the low sample contained endogenous CRP and IP-10 concentrations. The high and low samples were mixed at varying proportions to assay the proposed linear range.

### Whole Blood Precision

The precision of K2EDTA whole blood measurements was also estimated using fresh venous whole blood from five healthy adult donors. Two samples were derived from each donor; one contained endogenous levels of CRP and IP-10, while the other was spiked with 30 mg/L of CRP and 330 pg/mL of IP-10. Each sample was in six replicates on three cartridge lots across two days (a total of 30 replicates per sample). The coefficient of variation (%CV) was calculated for each sample.

### Matrix Equivalency

To broaden the assay implementation to whole blood, we quantified CRP and IP-10 using matched whole blood and plasma from healthy donors. Contrived samples were prepared by spiking analytes into whole blood (CRP at 50 mg/L and IP-10 at 300 pg/mL). A linear regression was fitted to the data upon observing a correlation between measurement in K2EDTA plasma and K2EDTA whole blood. We then generated a scale factor to account for hematocrit for each analyte using the slope of this linear regression to adjust the concentration of CRP and IP-10 analytes in whole blood to match those measured in plasma. We then tested matrix equivalency on 100 matched serum and whole blood samples from patients enrolled in our PROTECT-APT protocol (NCT04844541).^20^ Samples were collected in person at Johns Hopkins Hospital and delivered by courier within four hours to the ACESO laboratory. First, 100 µl of fresh EDTA whole blood was removed for testing, and the SST serum tubes were centrifuged at 1500 x g for 10 minutes. Then 100 µl of serum was tested on the assay. The whole blood measurement was multiplied by the hematocrit scale factor (CRP*1.5, IP-10*1.29), CRP and IP-10 concentrations were plotted, and Pearson correlations were calculated.

### Validation with Clinical Samples

93 SARS-CoV-2 (COVID-19) positive, 17 Influenza-like Illnesses (ILI), and five healthy close contacts were enrolled at Johns Hopkins Hospital under The Prophylaxis and Treatment of COVID-19 – Adaptive Platform Trial (PROTECT-APT).^20^ Subjects were enrolled as COVID-19 positive after confirmation of a positive RT-PCR test result within <10 days of enrollment or enrolled as ILI after meeting the World Health Organization ILI case definition: “*An acute respiratory illness with a measured temperature of ≥38 °C and cough, with onset within the past ten days*.”^21^ A nasopharyngeal swab was also taken at day 0 and tested on the BioFire® Respiratory 2.1 Panel. Subjects were split by BioFire result into COVID, ILI, or Healthy. If the BioFire was negative, then enrollment status based on earlier testing and symptoms was used. Both fresh (delivered by courier on the same day) and frozen (delivered on dry ice after aliquoting and storing at -80°C) SST serum were tested on the LightDeck CRP/IP-10 duplex assay. In addition, Flu-PRO questionnaires were administered on the same day as each blood collection to assess overall symptom severity.

### Data Analysis and Plotting

Data was loaded into R^22^ statistical environment to be analyzed. To compare the analyte measurements between blood and serum samples, the data was plotted as an x-y scatter with a line depicting a perfect correlation and a regression line generated using linear models (*lm*) functionality of the *ggplot2*^23^ package. To generate boxplots with points and assess statistical differences between the groups, the data was filtered to retain only measurements in the instrument’s dynamic range. CRP values > 1.3 mg/ml in serum and > 2.6 mg/ml in blood were retained, and for IP-10, all measurements were in the dynamic range. The healthy ranges for CRP^24^ and IP10 were identified in previously published work. Statistical comparisons were performed using the t-test function in R^22^. Points overlaying boxplots were plotted using the *ggbeeswarm*^25^ package. Figures were finalized in Adobe Illustrator.

## Results

### Cartridge and Multiplex Assay Configuration

The CRP/IP-10 duplex assay is a single-use cartridge designed to quantitate CRP and IP-10 from 100 µL of the sample [whole blood (K2 EDTA), plasma (K2 EDTA), or serum] in a rapid, simple one-step multiplexed immunoassay. The manufacturer provides lot-specific calibrations, alleviating the need for end-users to carry out any cartridge calibration in the field. The Analyzer automates all processing steps once the cartridge is inserted and reports the concentration of CRP (in mg/L) and IP-10 (in pg/mL) on the instrument screen at the end of the test (Figure 1A). The cartridge contains four capture spots per analyte and control spots (Figure 1B). In a competition assay, CRP in the sample competes for the binding site of the capture antibody on the surface and inhibits its interaction with AF647-labeled CRP from the detection pellet (Figure 1C). The fluorescence intensity measured on these spots is inversely correlated to the concentration of CRP in the sample. The concentration of IP-10 is measured using a sandwich assay, and the fluorescence intensity is directly related to the concentration of IP-10 in the sample. Specifically, IP-10 present in the sample binds to capture the antibody immobilized on the surface, enabling the binding of the detected antibody and the formation of a sandwich (Figure 1C). The fluorescence of immobilized Alexa Fluor 647 dyes at spots corresponding to each analyte is then measured with an integrated CMOS camera and interpreted by integrated software. Waveguide-mediated fluorescence only excites fluorophores captured on the surface within the evanescent field (∼100 nm) (Figure 1C). The LightDeck analysis software averages the fluorescence intensities of four capture spots and returns a single intensity value for each analyte.

### Evaluation of Multiplexed Quantification on the LightDeck Platform

The CRP/IP-10 duplex cartridge was calibrated using recombinant proteins spiked into the pooled human serum. To establish calibrated dose response, we used varying concentrations of each analyte. As a result, we observed the expected dose-dependent inverse correlation of signal to CRP concentration (competitive immunoassay) and the opposite, direct correlation between relative fluorescence units and concentration for IP-10 (sandwich immunoassay) (Figure 2). The CRP and IP-10 assays demonstrated high sensitivities with a Limit of Detection (LoD) of 0.6 mg/L and 12.8 pg/mL, respectively. The Limit of Quantitation (LoQ) was defined as two times the LoD, at 1.3 mg/L and 25.4 pg/mL, respectively. In addition, we observed a robust linear relationship between signal and analyte concentration for both CRP and IP-10 using whole blood samples containing different levels of analytes. To calculate percent recovery, we compared the mean of replicate values at each dilution against the expected value predicted by the dilution scheme. CRP/IP-10 duplex test showed linear results in K2EDTA whole blood, within 4.3 mg/L to 150 mg/L for CRP assay and 150 pg/mL up to 1,900 pg/mL for IP-10. Lastly, we evaluated assay precision using whole blood containing endogenous and spiked levels of CRP and IP-10. Samples were tested across different days, operators, instruments, and cartridge lots and demonstrated within-lab variation below 15%. These results confirm that our assay behaves as designed, is highly reproducible, and precise.

**Figure 2.**
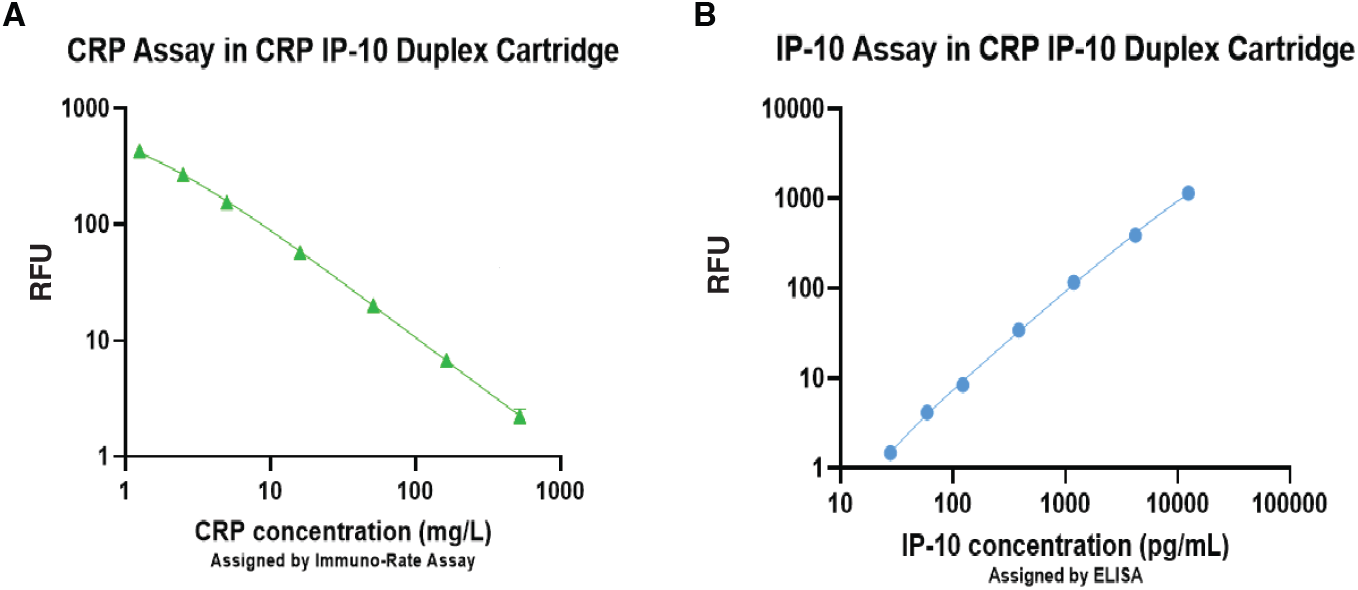
Analytical sensitivity of CRP IP-10 Duplex Assay. Dose-response curves of A) CRP (competitive immunoassay) and B) IP-10 (sandwich immunoassay) carried out with recombinant proteins spiked into pooled human serum. Relative fluorescence units (RFU) measure AF647 signal from the cartridge. Shown are mean of triplicate measurements with error bars. REPLOT

### Evaluation of Measurement Reproducibility in Blood and Serum

One of our goals was to design an assay that could be used at the point of care in resource-limited settings. For this purpose, whole blood is an optimal sample type for overcoming the equipment and time requirements of preparing plasma and serum. In addition, by testing whole blood directly, blood biomarkers can be quantified more rapidly and help clinicians make important diagnoses faster. We, therefore, set out to demonstrate equivalency between K2EDTA whole blood and serum matrices in the CRP/IP-10 duplex test. We tested matched samples from 100 subjects enrolled in the ongoing observational Prophylaxis and Treatment of COVID-19 – Adaptive Platform Trial (PROTECT-APT) study (NCT04844541).^20^ These samples were collected at different time points from COVID-19-positive outpatients, patients with Influenza-like illness (ILI), and healthy close contacts. Our measurements showed serum CRP values ranging from the assay limit of detection <1.3 to 94.2 mg/L and serum IP-10 values ranging from <25.4 to 5156.5 pg/ml. After excluding data points below the limit of detection (for CRP), we observed a high correlation for CRP (r = 0.993) and IP-10 (r = 0.924) between blood and serum after adjusting for matrix bias in whole blood measurements with a hematocrit scale factor (Figure 3). These results showed that CRP and IP-10 measurements in blood and serum are highly concordant and closely correlated.

**Figure 3.**
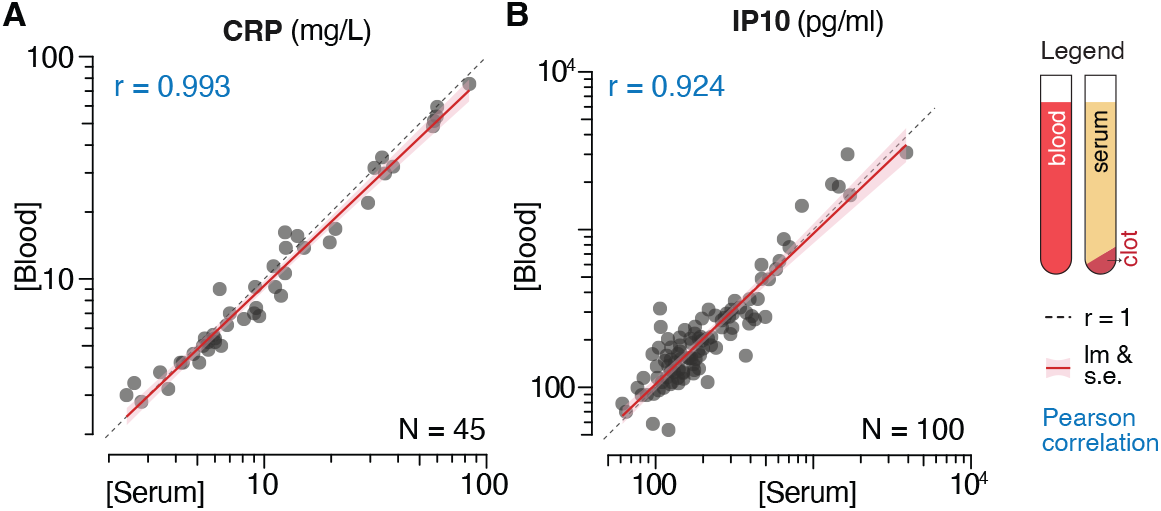
Blood and serum analyte measurements are highly concordant. Comparison of paired measurements of (A) CRP and (B) IP10 analytes between the whole blood and serum. Measurements of both analytes are highly correlated (see Pearson r values). The diagonal line indicates perfect correlation, and the colored line corresponds to linear regression. N indicates the number of measurements in the dynamic range. For CRP, 45 measurements were below the limit of quantification (LoQ) of 1.3 mg/L.

### Validating the CRP/IP-10 Duplex Test with Clinical Patient Samples

To validate our assay in a clinical context, we used clinical samples from the currently ongoing observational PROTECT-APT study. We collected 264 serum samples at multiple time points (day 0, 7, 14, 28) from COVID-19 outpatients, ILI patients, and healthy close contacts. Flu-PRO symptom questionnaires were collected on the same day as venipuncture and used to assess overall symptom severity.^26^ Subjects who did not complete the Flu-PRO questionnaire were excluded from the analysis. In addition, we segregated patients according to Flu-PRO self-scored symptom severity into four groups: “*No symptoms today*,” “*Mild*,” “*Moderate*,” or “*Severe*” symptoms. We found significant differences between CRP and IP-10 levels related to their overall symptom severity (Figure 4). The CRP^24^ and IP-10^27^ levels of subjects experiencing no symptoms most often fell within the healthy published ranges for these analytes. Importantly, subjects experiencing mild symptoms at the time of collection had significantly higher levels of CRP and IP-10 (p-value < 0.01), and those subjects experiencing moderate and severe symptoms also had significantly increased CRP and IP-10 levels compared to those with “*No symptoms*” (Figure 4, significance indicated with stars).

**Figure 4.**
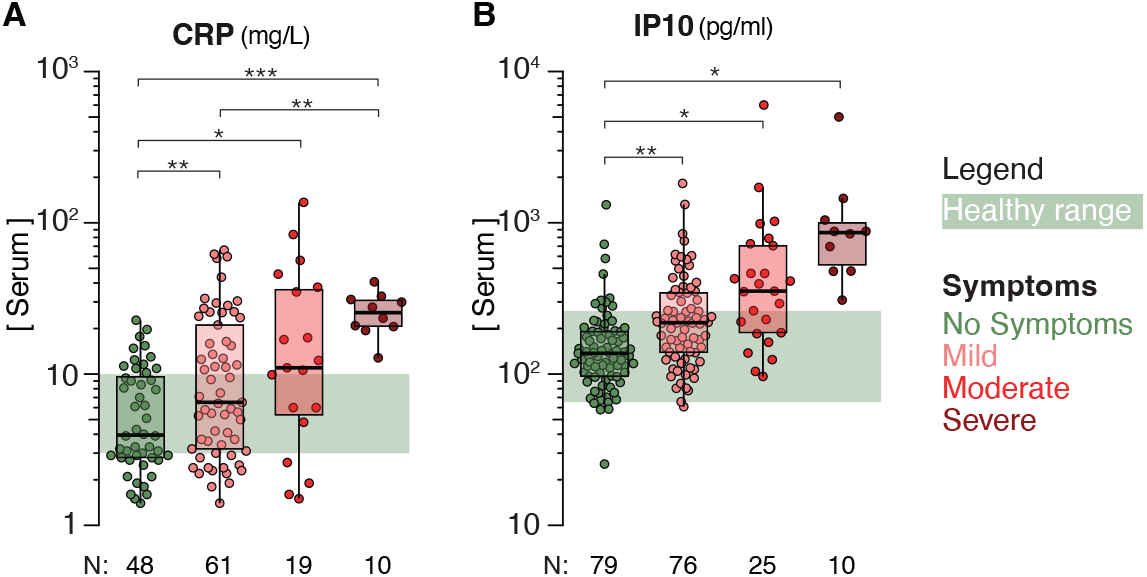
CRP and IP10 analyte levels and symptom severity. The measurements of (A) CRP and (B) IP10 analytes were grouped by the overall symptom severity. Boxplots and dots show measurements in dynamic range and a number of measurements (N) are shown below plots. For CRP, measurements below the limit of quantification (LoQ) are excluded. Significance was calculated with the Welch Two-Sample t-test (p.value ranges: * ≤ 0.05, ** ≤ 0.01,*** ≤ 0.001). The shaded green regions indicate the published analyte range in healthy subjects (CRP: 3-10 mg/L, IP10: 65-261 pg/ml).

Furthermore, we were able to discern differences between symptom severity groups (CRP: mild vs. severe) despite the limiting sample size (lowest N = 10) in multiple instances. Next, we compared the CRP and IP-10 levels between “*Healthy*” close contacts, COVID-19 positive, and patients with Influenza-like symptoms (ILI) (Figure 5). For the few healthy subjects, we observed analyte levels within healthy published ranges ^24,27^. Furthermore, CRP and IP-10 were both significantly elevated (p-value <0.001) in COVID-19 patients at day 0 of enrollment. IP-10 levels were broadly distributed for ILI subjects, with about half being elevated and the other half falling within the “healthy” range (Figure 5B). After performing the BioFire assay respiratory panel, we confirmed viral infection in half of the ILI subjects. These results suggest that the elevated IP-10 levels in half of the ILI patients could be due to elevated levels of IP-10 seen in viral infections. ^28–30^ Altogether, our results demonstrate the use of the Lighdeck CRP/IP-10 duplex assay to reproducibly measure CRP and IP-10 levels in blood or serum and exemplify how these measurements can be used to distinguish between healthy and infected subjects or to compare symptom severity in the context of the clinical study.

**Figure 5.**
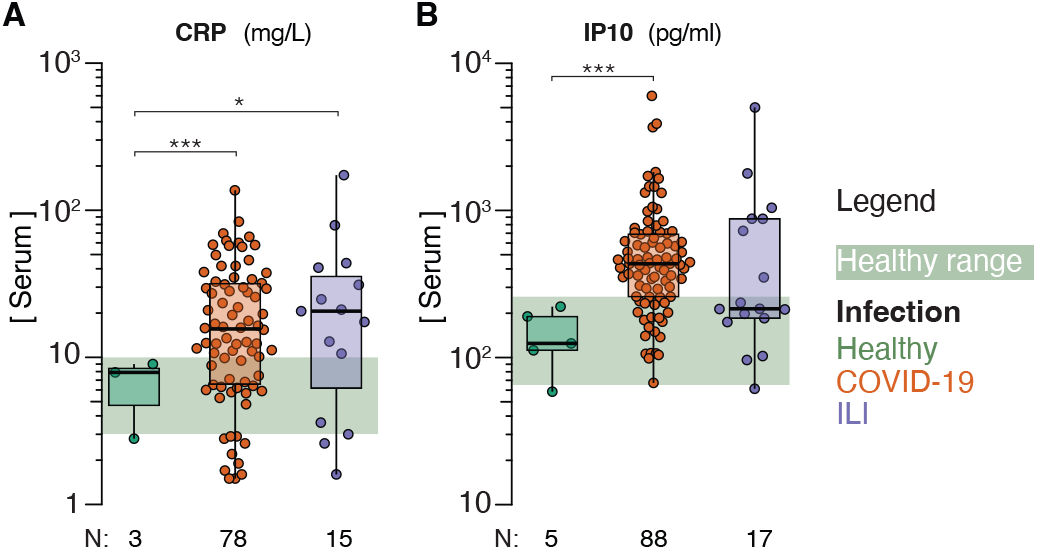
CRP and IP10 levels in healthy and infected subjects. The serum concentration of the (A) CRP and (B) IP10 at day 0 between healthy, COVID-19, and patients with influenza-like symptoms (ILI). Boxplots and dots show measurements in dynamic range and a number of measurements (N) are shown below plots. For CRP, measurements below the limit of quantitation (LoQ) are excluded. Significance was calculated with the Welch Two-Sample t-test (p.value ranges: * ≤ 0.05, ** ≤ 0.01,*** ≤ 0.001). The shaded green regions indicate the published analyte range in healthy subjects (CRP healthy range: 3-10 mg/L, IP10 healthy range: 65-261 pg/ml).

## Discussion

Clinical diagnoses are becoming increasingly complicated because of highly individualized immune responses to different pathogens and a rapidly growing number of biomarkers associated with each response. As a result, clinicians often encounter the diagnostic challenge of distinguishing between bacterial and viral etiologies in a febrile patient.^31^ Medical history, physical findings, and other ancillary medical tests are frequently similar for different causative agents and do not provide definitive discrimination.^32,33^ This could lead to misdiagnosis, alteration of patient care, and misuse of antibiotics, which can have significant consequences for individual and global health. Host-based diagnostic and prognostic tools, such as the CRP/IP-10 duplex assay described here, offer a promising solution to this increasingly complex problem. A transcriptomic assay measuring mRNA host response classifiers (including CRP and IP-10) to predict COVID-19 severity and 30-day outcome from the infection and the development of an electrochemical sensing platform for IL-10 or CRP are only a few examples of creative approaches being pursued to support more accurate medicine on an individual level.^12,13,34^ From among many biomarkers, IP-10 has gained prominence because it is elevated in a number of other viral infections, including Nipah virus^35^, and is elevated in the serum of patients with Rift Valley fever, chikungunya and severe H1N1 influenza A virus.^36–38^ In addition to being relevant to other diseases, including COVID-19, cancer, or Tuberculosis, the solid diagnostic and prognostic capabilities of IP-10 are most remarkable when measured alongside other biomarkers. ^12^ In fact, a host immune blood-based diagnostic proteomic signature integrating the concentrations of IP-10, CRP, and tumor necrosis factor-related apoptosis-induced ligand (TRAIL) has been shown to be able to distinguish bacterial from viral infection by multiple groups.^28–30^ These previously published studies and our work together provide strong support for the development of simple, rapid, cost-effective, and combinatorial biomarker measurement platforms as powerful tools to support clinical decision-making.

## Data Availability

All data produced in the present study are available upon reasonable request to the authors.

## Acknowledgments

We thank the PROTECT-APT study team at Johns Hopkins Hospital for their contribution to obtaining clinical samples.

## References

1. Osuchowski, M. F. et al. The COVID-19 puzzle: deciphering pathophysiology and phenotypes of a new disease entity. The Lancet Respiratory Medicine vol. 9 Preprint at https://doi.org/10.1016/S2213-2600(21)00218-6 (021).

2. Del Valle, D. M. et al. An inflammatory cytokine signature predicts COVID-19 severity and survival. Nat Med 26, (2020).

3. Malik, P. et al. Biomarkers and outcomes of COVID-19 hospitalisations: systematic review and meta-analysis. BMJ Evid Based Med 26,107 (2021).

4. Herold, T. et al. Elevated levels of IL-6 and CRP predict the need for mechanical ventilation in COVID-19. Journal of Allergy and Clinical Immunology 146, (2020).

5. Lev, S. et al. Observational cohort study of IP-10’s potential as a biomarker to aid in inflammation regulation within a clinical decision support protocol for patients with severe COVID-19. PLoS One 16, (2021).

6. Laing, A. G. et al. A dynamic COVID-19 immune signature includes associations with poor prognosis. Nat Med 26, (2020).

7. Rizzi, M. et al. Prognostic Markers in Hospitalized COVID-19 Patients: The Role of IP-10 and C-Reactive Protein. Dis Markers 2022, (2022).

8. Gudowska-Sawczuk, M. & Mroczko, B. What Is Currently Known about the Role of CXCL10 in SARS-CoV-2 Infection? International Journal of Molecular Sciences vol. 23 Preprint at https://doi.org/10.3390/ijms23073673 (2022).

9. Luan, Y. Y., Yin, C. H. & Yao, Y. M. Update Advances on C-Reactive Protein in COVID-19 and Other Viral Infections. Frontiers in Immunology vol. 12 Preprint at https://doi.org/10.3389/fimmu.2021.720363 (2021).

10. Blot, M. et al. CXCL10 could drive longer duration of mechanical ventilation during COVID-19 ARDS. Crit Care 24, (2020).

11. Yang, Y. et al. Plasma IP-10 and MCP-3 levels are highly associated with disease severity and predict the progression of COVID-19. Journal of Allergy and Clinical Immunology 146, (2020).

12. Madhurantakam, S. et al. Multiplex sensing of IL-10 and CRP towards predicting critical illness in COVID-19 infections. Biosens Bioelectron X 13, (2023).

13. Angel, A. et al. 32. Host Immune-Protein Signature Combining TRAIL, IP-10 and CRP for Early and Accurate Prediction of Severe COVID-19 Outcome. Open Forum Infect Dis 8, (2021).

14. Logan, C. et al. Rapid multiplexed immunoassay for detection of antibodies to Kaposi’s sarcoma-associated herpesvirus. PLoS One 11, (2016).

15. Broger, T. et al. Diagnostic performance of tuberculosis-specific IgG antibody profiles in patients with presumptive tuberculosis from two continents. Clinical Infectious Diseases 64, (2017).

16. Reverté, L. et al. Tetrodotoxin detection in puffer fish by a sensitive planar waveguide immunosensor. Sens Actuators B Chem 253, (2017).

17. Bickman, S. R. et al. An Innovative Portable Biosensor System for the Rapid Detection of Freshwater Cyanobacterial Algal Bloom Toxins. Environ Sci Technol 52,11691–11698 (2018).

18. Lochhead, M. J. et al. Rapid multiplexed immunoassay for simultaneous serodiagnosis of HIV-1 and coinfections. J Clin Microbiol 49, (2011).

19. CLSI EP17-A2. Evaluation of detection capability for clinical laboratory measurement procedures. Wayne PAL Clinical and Laboratory Standards institute vol. 32 (2012).

20. Henry M. Jackson Foundation for the Advancement of Military Medicine. Prophylaxis and Treatment of COVID-19 (PROTECT-APT). National Library of Medicine (U.S) Preprint at https://clinicaltrials.gov/ct2/show/NCT04844541 (2021).

21. Fitzner, J. et al. Revision of clinical case definitions: Influenza-like illness and severe acute respiratory infection. Bull World Health Organ 96, (2018).

22. R Core Team. R: A Language and Environment for Statistical Computing. Preprint at https://www.R-project.org/ (2022).

23. Wickham, H. ggplot2 - Elegant Graphics for Data Analysis (2nd Edition). Journal of Statistical Software vol. 77 (2017).

24. Nehring, S. M., Goyal, A. & Patel, B. C. C Reactive Protein. in StatPearls [Internet] (StatPearls Publishing, 023).

25. Erik Clarke and Scott Sherrill-Mix and Charlotte Dawson. ggbeeswarm: Categorical Scatter (Violin Point) Plots. Preprint at https://CRAN.R-project.org/package=ggbeeswarm (2022).

26. Richard, S. A. et al. Performance of the inFLUenza Patient-Reported Outcome Plus (FLU-PRO Plus) Instrument in Patients With Coronavirus Disease 2019. Open Forum Infect Dis 8, (2021).

27. Gotsch, F. et al. CXCL10/IP-10: A missing link between inflammation and antiangiogenesis in preeclampsia? Journal of Maternal-Fetal and Neonatal Medicine 20, (2007).

28. Oved, K. et al. A novel host-proteome signature for distinguishing between acute bacterial and viral infections. PLoS One 10, (2015).

29. van Houten, C. B. et al. A host-protein based assay to differentiate between bacterial and viral infections in preschool children (OPPORTUNITY): a double-blind, multicentre, validation study. Lancet Infect Dis 17, (2017).

30. Ashkenazi-Hoffnung, L. et al. A host-protein signature is superior to other biomarkers for differentiating between bacterial and viral disease in patients with respiratory infection and fever without source: a prospective observational study. European Journal of Clinical Microbiology and Infectious Diseases 37, (2018).

31. Baron, E. J. et al. Executive Summary: A Guide to Utilization of the Microbiology Laboratory for Diagnosis of Infectious Diseases: 2013 Recommendations by the Infectious Diseases Society of America (IDSA) and the American Society for Microbiology (ASM)a. Clinical Infectious Diseases 57, (2013).

32. Van Den Bruel, A. et al. Diagnostic value of laboratory tests in identifying serious infections in febrile children: Systematic review. BMJ 342, (2011).

33. Craig, J. C. et al. The accuracy of clinical symptoms and signs for the diagnosis of serious bacterial infection in young febrile children: Prospective cohort study of 15 781 febrile illnesses. BMJ (Online) 340,(2010).

34. Nikhil, R.-M. et al. Using a 29-mRNA Host Response Classifier To Detect Bacterial Coinfections and Predict Outcomes in COVID-19 Patients Presenting to the Emergency Department. Microbiol Spectr 10,e02305.22 (2022).

35. Mathieu, C. et al. Lethal nipah virus infection induces rapid overexpression of cxcl10. PLoS One 7,p (2012).

36. Jansen Van Vuren, P. et al. Serum levels of inflammatory cytokines in Rift Valley fever patients are indicative of severe disease Emerging viruses. Virol J 12, (2015).

37. Venugopalan, A., Ghorpade, R. P. & Chopra, A. Cytokines in acute chikungunya. PLoS One 9,p (2014).

38. Hagau, N. et al. Clinical aspects and cytokine response in severe H1N1 influenza A virus infection. Crit Care 13, (2010).

